# Sensitivity analysis of the effects of non-pharmaceutical interventions on COVID-19 in Europe

**DOI:** 10.1101/2020.06.15.20131953

**Authors:** Kristian Soltesz, Fredrik Gustafsson, Toomas Timpka, Joakim Jaldén, Carl Jidling, Albin Heimerson, Thomas B. Schön, Armin Spreco, Joakim Ekberg, Örjan Dahlström, Fredrik Bagge Carlson, Anna Jöud, Bo Bernhardsson

**Affiliations:** Lund University, Department of Automatic Control, Sweden; Linköping University, Department of Electrical Engineering, Sweden; Linköping University, Department of Health, Medicine and Caring Sciences, Sweden; Region Östergötland, Unit for Public Health and Statistics, Sweden; KTH Royal Institute of Technology, Division of Information Science and Engineering, Sweden; Uppsala University, Department of Information Technology, Sweden; Linköping University, Department of Behavioural Sciences and Learning, Sweden; National University of Singapore, Acoustic Research Laboratory, Singapore; Lund University, Department of Laboratory Medicine, Sweden; Skåne University Hospital, Department of Research and Development, Lund, Sweden

**Author notes:** **Author’s contributions** Study conceptualisation: Bernhardsson, Gustafsson, Jaldén, Soltesz, Timpka. Analysis and interpretation of model weaknesses: Bernhardsson, Bagge Carlson, Jaldén, Soltesz Code preparation and execution: Heimerson, Jidling. Background literature review: Bernhardsson, Jöud, Spreco,Timpka. Manuscript writing and reviewing: Bernhardsson, Dahlström, Ekberg, Jaldén, Jöud, Schön, Soltesz, Spreco, Timpka. **Competing interests**: None.

## Abstract

The role of non-pharmaceutical interventions (NPIs) on the spread of SARS-CoV-2 has drawn significant attention, both scientific and political. Particularly, an article by the Imperial College COVID-19 Response Team (ICCRT), published online in Nature on June 8, 2020, evaluates the efficiency of 5 NPIs. Based on mortality data up to early May, it concludes that only one of the interventions, lockdown, has been efficient in 10 out of 11 studied European countries.

We show, via simulations using the ICCRT model code, that conclusions regarding the effectiveness of individual NPIs are not justified. Our analysis focuses on the 11th country, Sweden, an outlier in that no lockdown was effectuated. The new simulations show that estimated NPI efficiencies across all 11 countries change drastically unless the model is adapted to give the Swedish data special treatment. While stated otherwise in the Nature article, such adaptation has been done in the model code reproducing its results: An ungrounded country-specific parameter said to have been introduced in all 11 countries, is in the code only activated for Sweden. This parameter *de facto* provides a new NPI category, only present in Sweden, and with an impact comparable to that of a lockdown.

While the considered NPIs have unarguably contributed to reduce virus spread, our analysis reveals that their individual efficiency cannot be reliably quantified by the ICCRT model, provided mortality data up to early May.

## Introduction

The spread of SARS-CoV-2 has challenged the world and political decisions are continuously made to reduce the virus’s effect on population health and on healthcare organisations. In the absence of a vaccine, populations can only safeguard themselves through non-pharmaceutical measures ranging from increased personal hygiene to social distancing. Different non-pharmaceutical interventions (NPIs) have been effectuated across the world. Scientific guidance is therefore needed upon which to base national exit strategies from implemented NPIs in a safe, yet efficient, manner.

A series of reports have presented estimated effects of various NPIs on the spread of SARS-CoV-2 infection, such as case isolation, closure of schools, restrictions specifically towards vulnerable groups such as people above the age of 70, or a total lockdown. Specifically, two reports by the *Imperial College COVID-19 Response Team* (ICCRT) set out to quantify the effect of NPIs [1,2]. These reports have since been widely used as support for political decisions that aim at reducing the rate at which the virus spreads [3].

The first of these, *Report 9* [1], released online March 16, 2020, was influential in the change of the COVID-19 response policy in the UK [3] and is likely to have affected policy decisions in other European countries.

In the second one, *Report 13* [2], released online on March 30, a method was presented to estimate the actual effects of different NPIs that at the time had been effectuated in 11 European countries. Based on the effectuation dates of the interventions and available time series of mortalities in each country, the influence of individual NPIs on the reproduction number *R*_*t*_ of SARS-CoV-2 infection was estimated from death data.

Data from different countries were effectively pooled through the assumption that the NPI parameters are not country-specific: the factor of relative change in *R*_*t*_ resulting from effectuating a particular NPI is the same, regardless of in which country the NPI is effectuated. The factors were assumed time-independent, with the exception that a bonus factor is ascribed to an NPI if it is the first to be effectuated in a country. Some country-specific flexibility was, however, provided through the basic reproduction number *R*_*0*_. In a later version of the model [9], released April 24 (after Report 13), the lockdown NPI, being the last NPI to be effectuated in all countries, except for Sweden, was also ascribed some country-specific flexibility.

A slightly modified version of *Report 13* [2] was accepted for publication in Nature on May 22 and published online as an “accelerated article preview” on June 8 [4]. The Nature publication was based on a version of the method used in [2], with some subtle yet important modifications. The most notable such modification, discussed further below, was the introduction of a country-specific parameter to model the efficiency of the last NPI to be introduced in a country.

It is claimed in [4] that “in our analysis we find that only the effect of lockdown is identifiable”, but analysis supporting this claim is not to be found in [4], nor in [2] or [9].

The modellers behind [2,4] have openly shared their method implementation—a Bayesian estimation model—and data, through an online version-tracking repository [9]. This has enabled the reproduction of results from [2], [4], and, more importantly, analyses of model sensitivities and parameter identifiability. Such analyses, routinely performed in fields of the engineering sciences when estimation and prediction models are considered, have been largely omitted in [2] and [4]. While sensitivity analyses alone cannot ascertain the reliability of a model candidate, they can reveal the opposite.

## Objectives

In this study, we investigate the method used by the ICCRT [4] to estimate the effect of non-pharmaceutical interventions on the reproduction of SARS-CoV-2 in Europe. In particular, we examine model sensitivities and how problems with parameter identifiability influence the reliability of the results. A similar investigation of the model used by the ICCRT in their *Report 13* [2] is available in our previous report [5].

## Estimating the impact of interventions

In [4], five NPI categories are defined, as shown in Figure 1. When effectuated, each intervention is assumed to instantaneously result in a step-change in the reproduction number *R*_*t*_, as illustrated in Figure 2.

**Figure 1.**
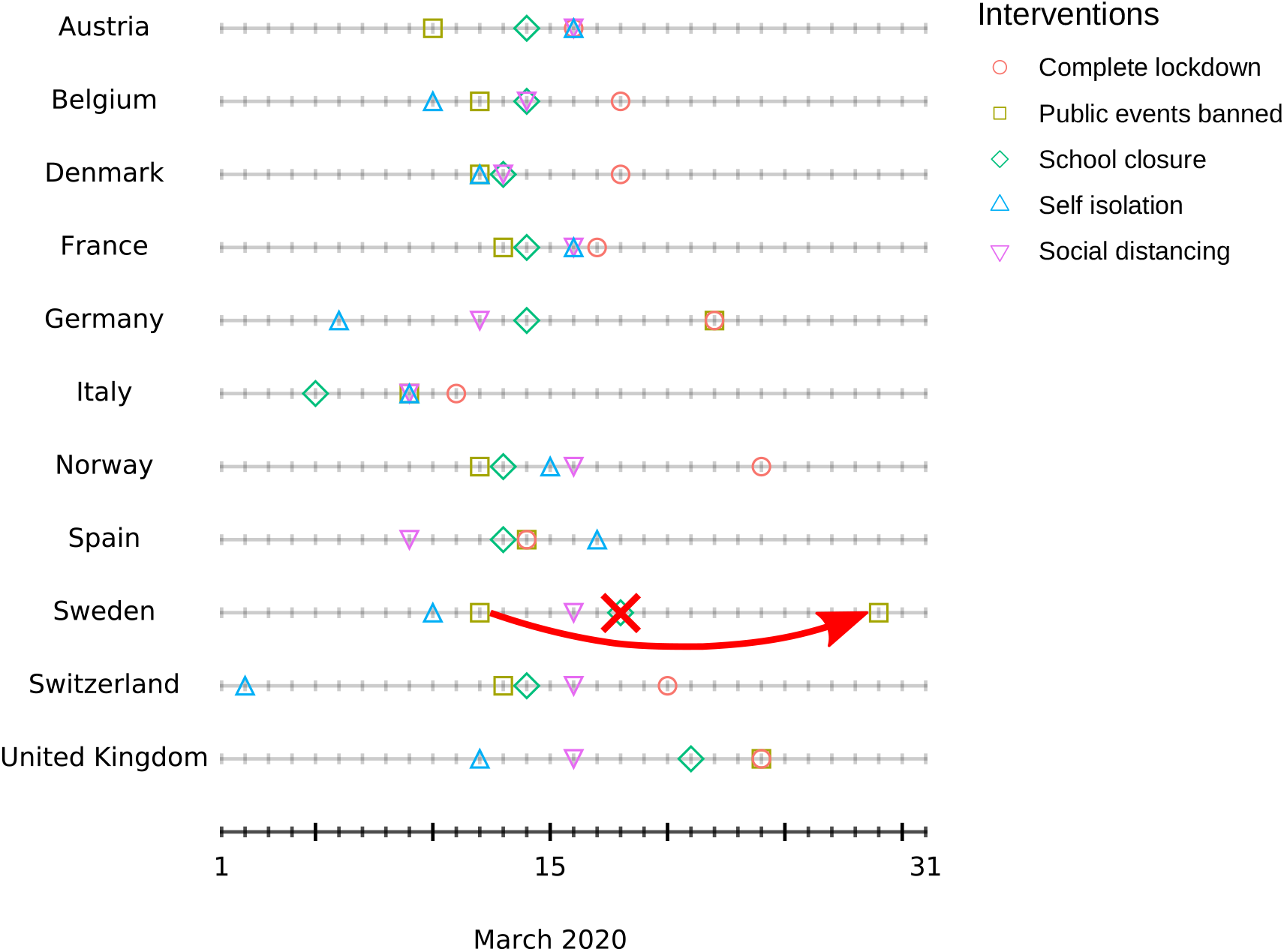
Dates that non-pharmaceutical interventions (NPIs) were effectuated in individual countries, according to [4]. The annotations illustrate code revisions introduced by the authors of [4] in their model code [9]: On April 9, the date on which *public events* were banned in Sweden was revised from March 12 to March 29. In the same code revision, *school closure* was no longer defined to have taken place in Sweden. In [4], *school closure* had again been defined to have occurred in Sweden. However, this does not comply with the corresponding version of the code [9], in which *school closure* remains defined as not have taken place in Sweden.

**Figure 2.**
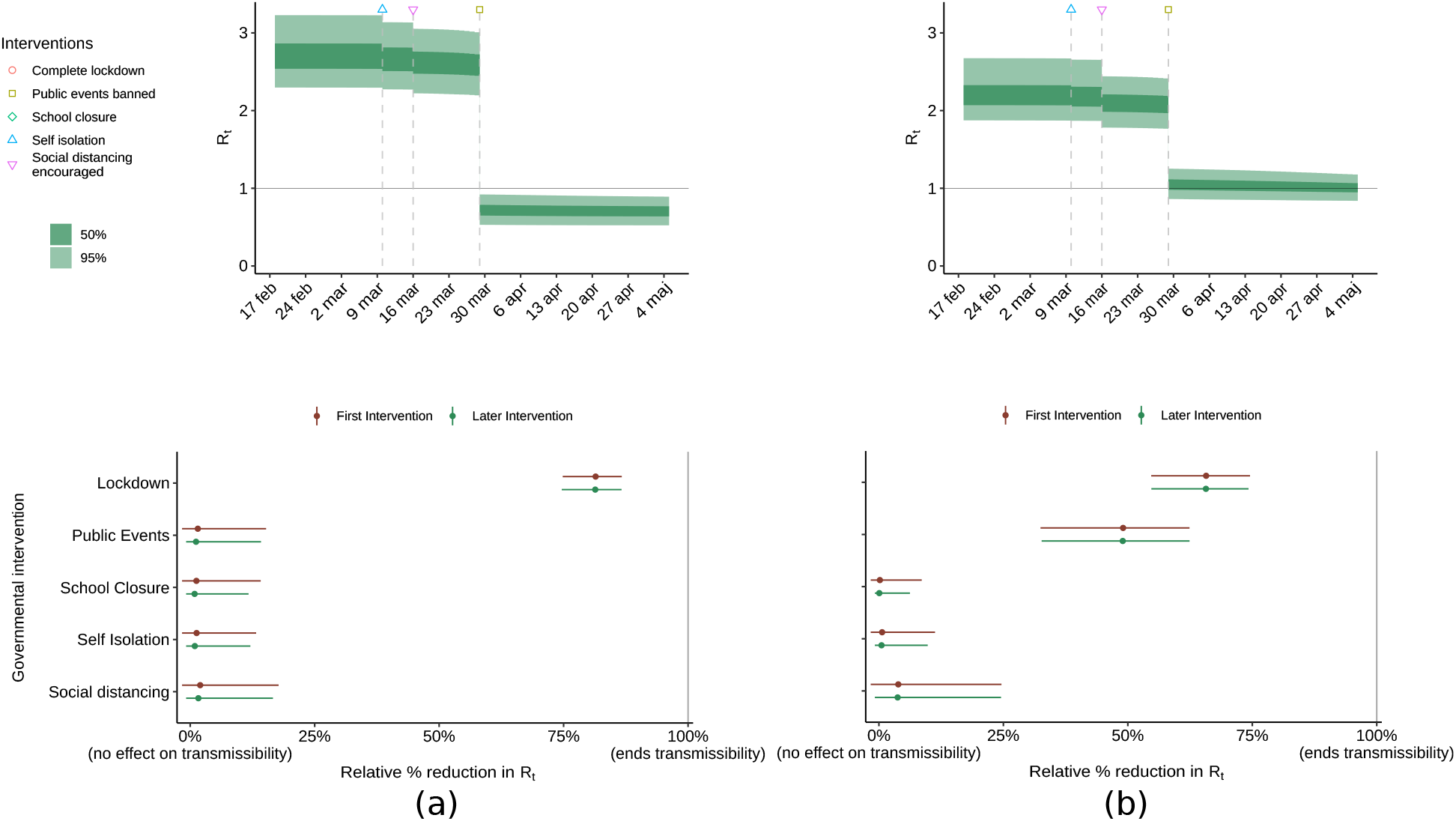
Estimated effectiveness of the *public events* ban intervention in Sweden. Top pane shows posterior credibility intervals for the reproduction number *R*_*t*_ in Sweden; the bottom pane shows the effectiveness of the considered interventions in the 11 modelled European countries. (a) Results of the code used in [4], with a separate last intervention parameter for the *public events* ban in Sweden. (b) Results of the same code, but with the last intervention parameter fix disabled also for Sweden, as in [2].

The model assumes that the reproduction number *R*_*t*_ is only affected by the NPI parameters, and by herd-immunity effects resulting from a diminishing susceptible-to-infected ratio. A partial data pooling is performed, in the sense that a particular NPI is expected to have the same multiplicative impact on *R*_*t*_, regardless of which country it is effectuated in. Three exceptions apply: I: The first NPI to be effectuated in a particular country is ascribed a bonus effect, as shown by the red *versus* green bars in Figure 2; II: lockdown is ascribed a country-specific bonus effect; III: the last NPI that is effectuated in a country, being the lockdown NPI for all countries except for Sweden, is also ascribed a country-specific bonus effect. Additional country-specific flexibility is provided by letting the basic reproduction number *R*_*0*_, be a country-specific parameter, co-identified with the NPI-effect parameters.

Apart from the NPI effectuation dates, a stochastic infection-to-death distribution and an infection-fatality-ratio are utilized. They are both based on previous reports [6]. Notably, the infection-to-death distribution (excluding reporting delay) has a mean of 23.9 days, comparable to the duration of the time window within which all NPIs were effectuated, as shown in Figure 1.

The above components are combined into a model, driven by country-wise daily reported deaths (source: ECDC [7]). A Bayesian approach based on Markov Chain Monte Carlo (MCMC) [8] simulations is employed to compute posterior distributions of the NPI effects on *R*_*t*_, along with the country-specific *R*_*0*_ values. In [4] the estimates are reported as 50 % and 95 % credibility intervals (CI) of these distributions.

It is claimed in [4] that lockdown has an “identifiable large impact on transmission” and that the “close spacing of interventions in time […] mean the individual effects of the other interventions are not identifiable”. Posterior credibility intervals for the effects of interventions presented in [4] however indicate the contrary: Individual 95 % CIs are presented for the effectiveness of each of the five considered NPIs. These CIs are all narrow, with each NPI except for lockdown ascribed effectiveness between -2 % and 18 %, see Figure 2 in [4].

## Methods and results

We performed an empirical analysis of the model underlying [4], using the original model code and input data as provided by the authors of [4] through [9]. This simulation-based analysis is specifically influenced by the case of one country that has received remarkable special treatment in the model: Sweden, the only country which has not implemented the *lockdown*.

### The role of the last intervention

While it was never motivated how data supports the exception of the lockdown, but not the other 4 NPIs from the otherwise applied pooling strategy introduced in [2], this special treatment of *lockdown* remained in [4].

As revealed in [5], the model version used in [2] had difficulties fitting the model to data from Sweden, which is the only modelled country in which *lockdown* had not been effectuated. This was fixed in the code version used in [4] by allowing country-specific variations of not only the lockdown intervention effect but also of “the last intervention to be effectuated in a country”. Notably, this fix was only activated for Sweden in the version of the code [9] reproducing the results of [4]. I.e., the fix introduces an ungrounded virtual NPI category for Sweden alone, that the model can utilise to improve its fit to data. When activating the last intervention fix for all countries, as is incorrectly claimed was done in [4], the statistics engine throws warnings that the “posterior means and medians may be unreliable”, indicating numerical issues.

With the fix in place for Sweden, the *public events* ban is ascribed a 71 % mean (95 % CI: 59– 81 %) reduction of *R*_*t*_ in Sweden, to be compared with its negligible < 2 % mean (and < 15 % with 95 % credibility) effect in all other modelled countries. Figure 2 shows the outcome for Sweden when executing the model with (a) and without (b) the last intervention fix. It should be noted that the removal of the model’s special treatment of Sweden in (b) significantly increases the estimated importance of the *public events* ban in all 11 countries, not just in Sweden.

We have not come across any reason motivating separate treatment of the last effectuated NPI, except for the one presented above, to also fit the Swedish data.

### Intervention definition sensitivity

A challenge in the modelling of NPI effects is to accurately represent when, and to what extent, a particular NPI was effectuated. We investigate two examples:

- Higher education instances in Sweden transitioned to online teaching on March 18, while elementary schools remained open. The version of the model used in *Report 13* [2] (dated March 30), defined *school closure* to have taken place on March 18. However, the model was revised on April 9, and *school closure* in Sweden was re-defined not to have taken place. In [4] school closure is again said to have been effectuated in Sweden, but in the code used to generate the results of [4] school closure is defined not to have taken place in Sweden.
- Similarly, public events exceeding 500 persons were banned in Sweden on March 12. On March 29, the restriction was tightened to 50 persons. *Report 13* defined the date of the *public events* ban to March 12. On April 9, the effectuation date of *public events* ban was changed from March 12 to March 29 in the code [9]. In [4], the effectuation date is defined to be March 29.

The outcome in Figure 3a is obtained when running the [4] version of the model, with school closure defined to have taken place in Sweden, and the Swedish *public events* ban to have taken place on March 29. With the *public events* ban date moved back to March 12, the results are instead those shown in Figure 3b. Re-running the model once more, now with *school closure* being defined not to have been effectuated, instead results in the estimates of Figure 3c. All parameters not explicitly mentioned above are kept at their nominal values used in [4].

**Figure 3.**
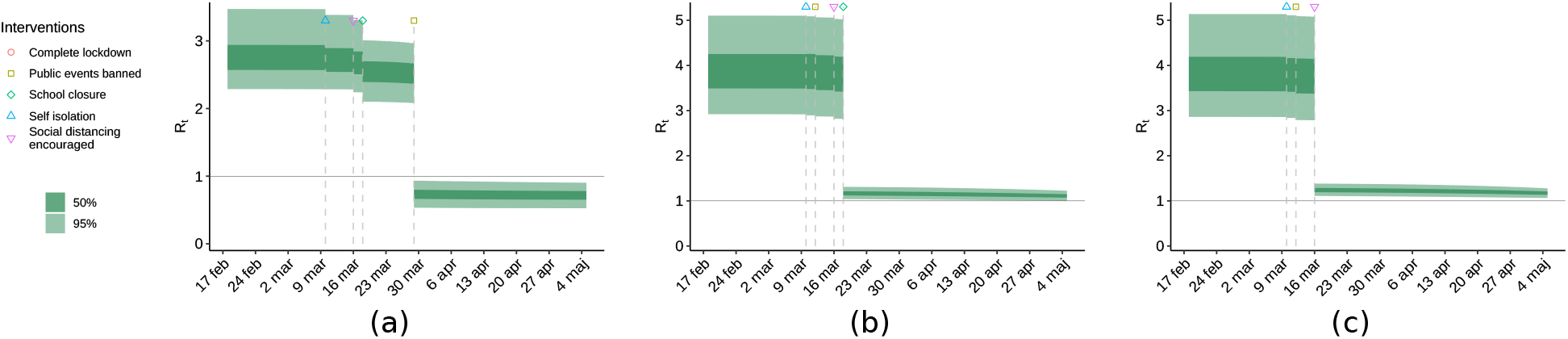
Effect of interventions on virus spread in Sweden, with slightly varying definitions of the interventions. **(a)** *School closure* defined to have taken place, *public events* defined to have been banned on March 29. **(b)** Same as (a), but with the *public events* ban moved back to March 12. **(c)** Same as (b), but with *school closure* defined not to have taken place. As expected, the visual appearance of the plots is similar, with the last intervention contributing most to the reduction of virus spread. This is problematic, since the last intervention differs between (a), (b), and (c), each relying on equally motivated NPI effectuation dates, de facto introduced by the authors of [2,4] in different versions of the model code [9].

As seen in Figure 3, subtle changes in NPI definitions have an unrealistically large impact on the estimated effectiveness of individual NPIs in Sweden. This is because the last effectuated NPI is exempt from the country data pooling and is hence the only NPI practically available to the model in order to explain the decrease in *R*_*t*_ indicated by the Swedish data in absence of a societal *lockdown*. The three scenarios attribute the most significant decrease in *R*_*t*_ to the *public events* ban, the *school closure*, and to *social distancing*, respectively.

Before the modellers, for undisclosed reasons, began treating the last effectuated NPI in each country separately, the above changes did not only impact the estimated effectiveness of *school closure* and *public events* ban in Sweden, but in all modelled countries, as further illustrated in [5]. This provides a second plausible explanation as to why the authors of [4] introduced the special treatment of the last NPI to be effectuated in each country.

### Estimating the importance of lockdown

With 11 countries in the analysis, Sweden was the only country without the NPI *lockdown* being effectuated. To give the model a chance to evaluate the efficiency of the *lockdown* intervention on a more equal footing, we ran the code on input data from only two countries: The UK and Sweden. We also deactivated the fix, present in [4] but not [2], that effectively exempts the last effectuated NPI in each country from the data pooling.

Pooling Sweden and UK data result in major reattributions of NPI effects, as shown in Figures 4 and 5. The model now attributes the reduction in *R*_*t*_ to the *public events* ban and estimates a negligible effect of the *lockdown* NPI.

**Figure 4.**
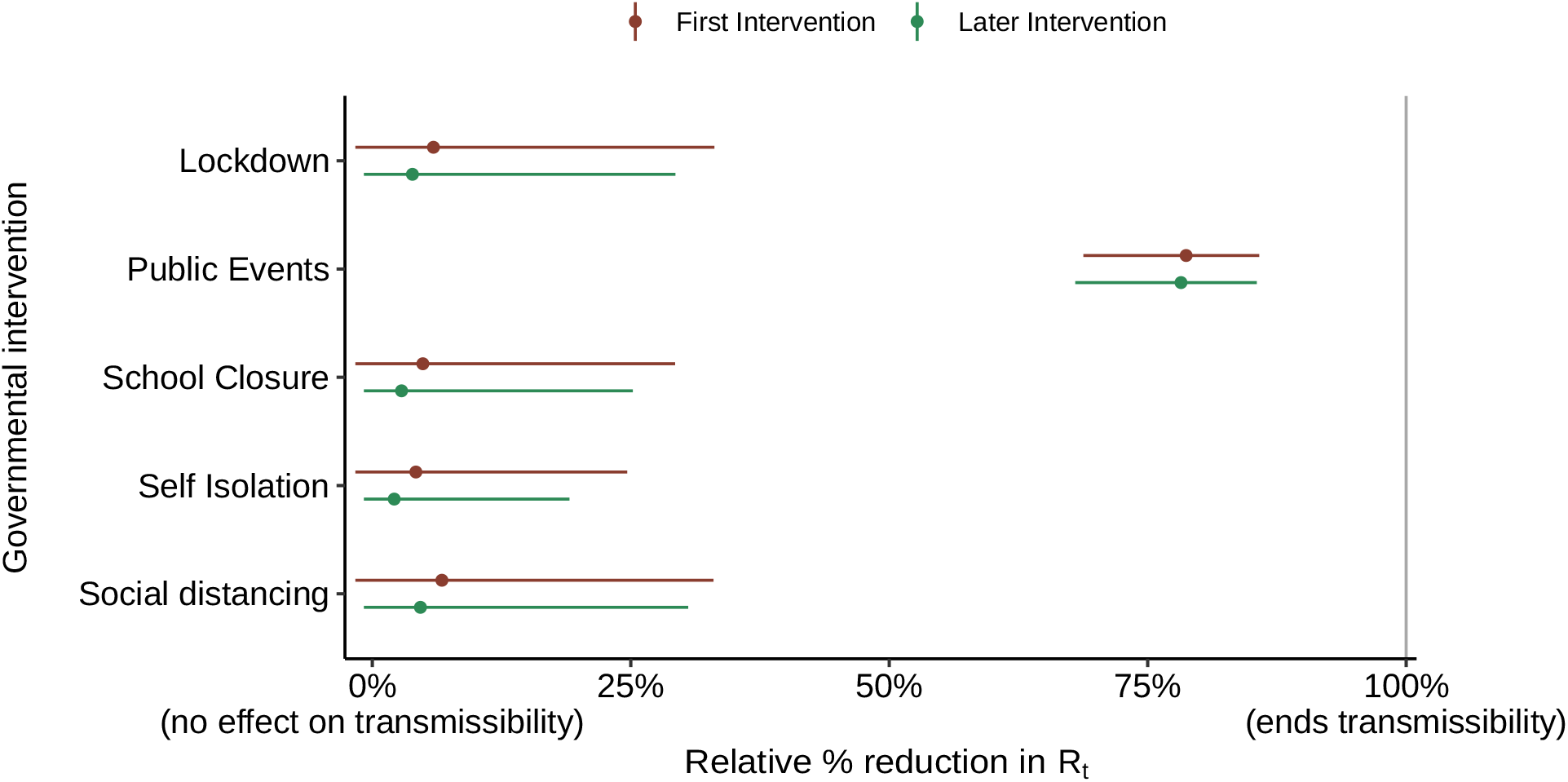
The estimated effectiveness of interventions. Results were obtained by pooling only two of the modelled countries: Sweden and the UK in an execution of the [4] version of the code [9], with the last intervention-fix disabled. The model now estimates that *public events* ban has been the most efficient intervention, while the importance of *lockdown* being small.

**Figure 5.**
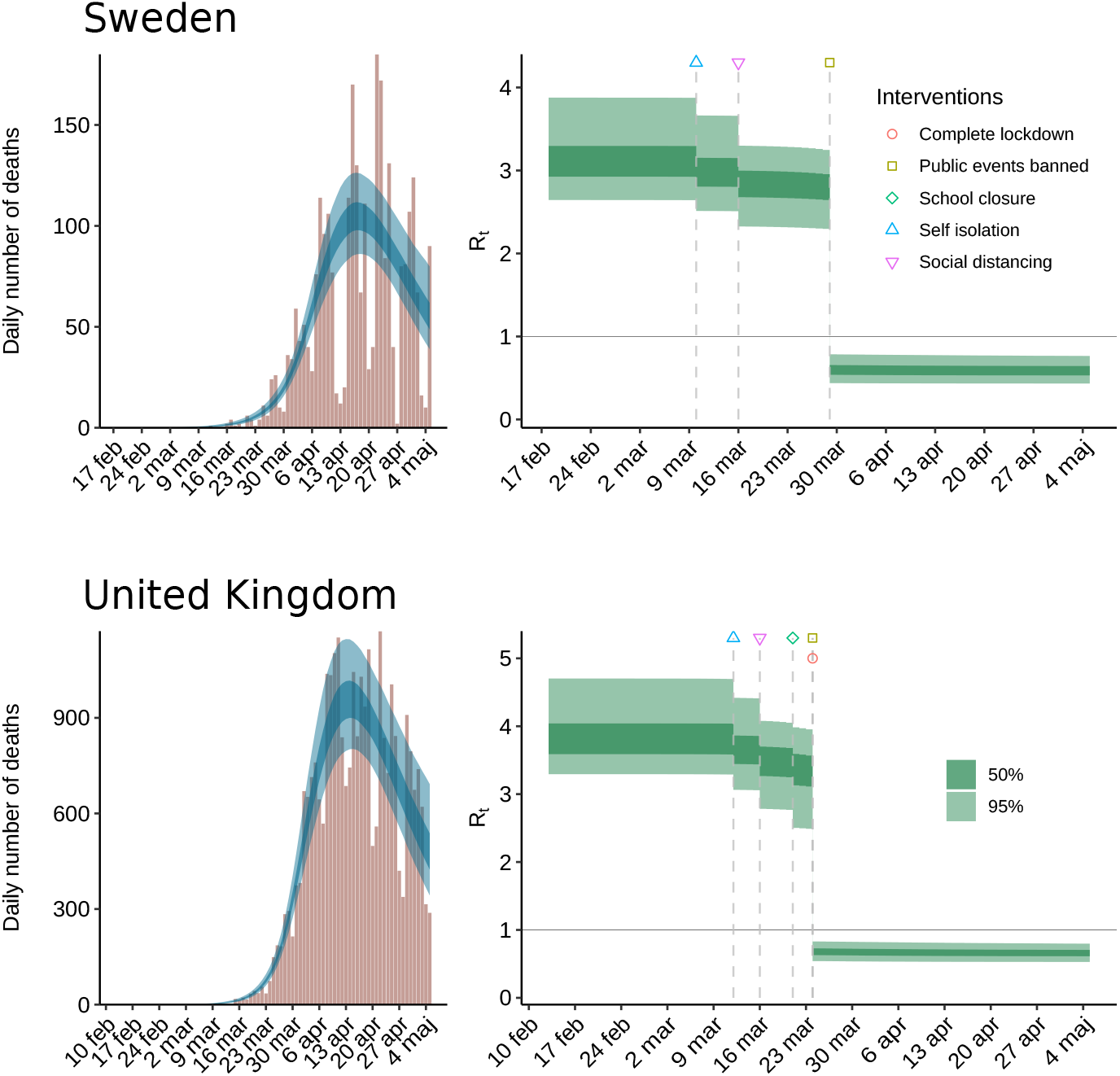
Estimated effectiveness of interventions. Results were obtained by pooling only two of the modelled countries: Sweden and the UK in an execution of the [4] version of the code [9], with the last intervention fix disabled. The reduction of *R*_*t*_ is almost exclusively ascribed to the *public events* ban; the model estimates the importance of *lockdown* to be negligible. The model estimates *R*_*t*_ to have dropped below 1 at the end of March in both countries, with stated credibility exceeding 95 %.

### Data and code availability

We have used the original model code and associated data, provided in [9] and described as “the exact code that was used” in [4]. A fork of this code, generating the figures in this manuscript, is provided in [10].

## Discussion

The aim of this study was to investigate the method used by the ICCRT in [4] for estimation of NPI effects on SARS-CoV-2 spread. We found the combination of high model sensitivity, and the assumption of *R*_*t*_ being driven solely by the NPIs, to constitute a fundamental limitation. This should be considered when the modelling results are used as a basis for policymaking.

The perhaps most remarkable result in [4] is that almost all the decrease of viral reproduction was attributed to the *lockdown* intervention in the 10 countries where it was effectuated. In those same 10 countries, the effects of the 5 other considered intervention categories were almost negligible. In the one country where *lockdown* was not effectuated (Sweden), it was instead the public events ban intervention that almost alone contributed to attaining *R*_*t*_<1. It seems unlikely that this would be the result of fortunate circumstances, where *lockdown* was implemented in exactly the 10 countries where it would have a large impact on *R*_*t*_, and omitted in the single country, where it was not needed. The distribution modelling the infection-to-death delay in [4] has a mean of 23.9 days, and so the model could not predict this outcome at the time most countries chose to effectuate *lockdown*, while one country did not.

An arguably more plausible explanation of the findings is that it is the last of all interventions, that in the model will contribute to bringing *R*_*t*_<1 in a specific country. This is confirmed by executing the model [9] with different interventions being defined as having occurred last in the one country where lockdown was not effectuated. It is hard to judge whether, for example, a partial transition to online teaching constitutes a *school closure* or not. This is reflected by the model used in [2] first defining *school closure* to have taken place in Sweden. In a subsequent code version it was defined not to have taken place, using the above motivation, and in the code version used in [4] it had again been re-introduced. Similarly, the crowd size limit associated with the *public events* ban NPI remains a free design parameter for the modeller to decide. Early versions of the model code defined the *public events* ban to have taken place on the date where gatherings exceeding 500 persons were discouraged in Sweden. This was later changed to the date when gatherings exceeding 50 persons were discouraged. As shown in Figure 1, the intervention defined to have been effectuated last in Sweden is different, depending on which of the mentioned definitions of *school closure* and *public events* ban is chosen. This results in either *school closure, public events*, or *social distancing discouraged* becoming defined as the last intervention to have been effectuated in Sweden. In each case, the model uses the last intervention to explain the reduction in *R*_*t*_ that becomes evident in the death reports collected over April and May.

The main difference between the models used by the ICCRT in [2] and [4] is the introduction of country-specific effect parameters for the last NPI to be effectuated. This last NPI is *lockdown* in all countries except Sweden, where it is *public events* ban, *school closure* or *social distancing encouraged*, depending on which version of NPI effectuation dates is adopted. This bonus parameter provides an explanation of the decrease of *R*_*t*_ in Sweden, despite the absence of *lockdown*. Remarkably, in the ICCRT model code [9] reproducing the results in the Nature article [4], the bonus last intervention parameter has been disabled for all countries except Sweden, de facto providing a bonus NPI parameter in Sweden for the model to use. Enabling the bonus parameter for all 11 countries, to match the wording in [4], results in numerical issues and warnings from the MCMC engine that the posterior distributions should not be trusted.

Our point is not to argue whether a school closure took place or not in Sweden, what the most appropriate crowd size limit is, or whether *lockdown* should be ascribed a higher or lower effect on than for instance *public events* ban. Instead, our findings highlight a remarkably large sensitivity to such minor changes in input data.

The nominal NPI effectuation dates considered in [2], do not suffer from the pathological case of any NPI being effectuated on the same day across all 11 countries, see Figure 1, but the observed sensitivity issues can still be qualitatively understood from the analysis of the regression matrix for the linear regression problem at the heart of estimating the NPI parameters, as performed in [5,10]. The identifiability issues are to some extent acknowledged in [4], by recognizing that “The close spacing of interventions in time […] mean the individual effects of the other interventions are not identifiable”. The basis for this deduction is not presented in [4]. More remarkably, it is easily overshadowed by the subsequent presentation of credible intervals for the effects of the different NPIs, and the claim that “Lockdown has an identifiable large impact on transmission (81% [75% - 87%] reduction”. As seen in Figure 3, the mentioned credible intervals shrink as more data become available, hiding the fundamental identifiability problems of the underlying model, and giving the results a false sense of reliability.

There are several other aspects of the model in [4] that deserve scrutiny. The authors of [4] write that they “assess whether there is evidence that interventions have so far been successful at reducing *R*_*t*_ below 1” and later hint at an affirmative conclusion by writing that they “estimate large reductions in the reproduction number *R*_*t*_ in response to the combined non-pharmaceutical interventions”. We argue that it is questionable to determine the joint impact of NPIs on *R*_*t*_ using a model based on the assumptions that changes in *R*_*t*_ are *de facto* only driven by the NPIs. The authors seem to acknowledge this in their counterfactual analysis, by stating that “even in the absence of government interventions we would expect *R*_*t*_ to decrease and therefore would overestimate the deaths in the no-intervention mode”. Yet, no serious effort to falsify the underlying hypothesis that the NPIs are the main reason for the reduction in *R*_*t*_ is ever presented. The closest we could find was a section of the supplement of [4], corresponding to Section 8.4.4 of [2], that describes an attempt to assess this assumption using a form of Gaussian process prior null hypothesis. There is, however, no reference to quantitative results, and no code to perform this experiment has been provided in [9].

Using simple but insightful executions of the original model code, we have demonstrated that even a seemingly simplistic and data-driven phenomenological model can suffer from severe sensitivity and identifiability issues. With much at stake during all phases of a pandemic, we conclude that it is crucial to thoroughly scrutinize any SARS-CoV-2 estimation or prediction model, prior to considering its use as decision support in policymaking. Such scrutiny relies on modellers following the good practice used by the authors of [4], of sharing open source code.

We would finally like to state that we strongly support the ambition of [4], to use available data to estimate the efficiency of different NPIs. However, the fact that the NPIs were introduced so closely in time during some weeks in March, unfortunately, made identifiability of individual NPIs infeasible.

## Acknowledgements

We would like to acknowledge Ericsson Research, Lund Sweden, for hosting our model runs in their data centre.

